# Sequential appearance and isolation of a SARS-CoV-2 recombinant between two major SARS-CoV-2 variants in a chronically infected immunocompromised patient

**DOI:** 10.1101/2022.03.21.22272673

**Authors:** Emilie Burel, Philippe Colson, Jean-Christophe Lagier, Anthony Levasseur, Marielle Bedotto, Philippe Lavrard, Pierre-Edouard Fournier, Bernard La Scola, Didier Raoult

**Affiliations:** IHU Méditerranée Infection, 19-21 boulevard Jean Moulin, 13005 Marseille, France; Aix-Marseille Univ., Institut de Recherche pour le Développement (IRD), Microbes Evolution Phylogeny and Infections (MEPHI), 27 boulevard Jean Moulin, 13005 Marseille, France; Assistance Publique-Hôpitaux de Marseille (AP-HM), 264 rue Saint-Pierre, 13005 Marseille, France; Aix-Marseille Univ., Institut de Recherche pour le Développement (IRD), Vecteurs – Infections Tropicales et Méditerranéennes (VITROME), 27 boulevard Jean Moulin, 13005 Marseille, France

**Author notes:** **Corresponding author:** Didier Raoult, IHU Méditerranée Infection, 19-21 boulevard Jean Moulin, 13005 Marseille, France.; Bernard La Scola, IHU Méditerranée Infection, 19-21 boulevard Jean Moulin, 13005 Marseille, France. Contributed equally (Emilie Burel, Philippe Colson).

**Keywords:** SARS-CoV-2, variant, recombination, chronic infection, immunosuppression

## Abstract

Genetic recombination is a major evolutionary mechanism among RNA viruses, and it is common in coronaviruses, including those infecting humans. A few SARS-CoV-2 recombinants have been reported to date whose genome harbored combinations of mutations from different mutants or variants, but a single patient’s sample was analyzed, and the virus was not isolated. Here, we re-port the gradual creation of a hybrid genome of B.1.160 and Alpha variants in a lymphoma patient chronically infected for 14 months, and we isolated the recombinant virus. The hybrid genome was obtained by next-generation sequencing, and recombination sites were confirmed by PCR. This consisted of a parental B.1.160 backbone interspersed with two fragments, including the spike gene, from an Alpha variant. Analysis of seven sequential samples from the patient decoded the recombination steps, including the initial infection with a B.1.160 variant, then a concurrent infec-tion with this variant and an Alpha variant, the generation of hybrid genomes, and eventually the emergence of a predominant recombinant virus isolated at the end of the patient’s follow-up. This case exemplifies the recombination process of SARS-CoV-2 in real life, and it calls for intensifying genomic surveillance in patients coinfected with different SARS-CoV-2 variants, and more gener-ally with several RNA viruses, as this may lead to the creation of new viruses.

## INTRODUCTION

A major evolutionary mechanism of RNA viruses is genetic recombination [1,2]. Recombinations are extremely common in coronaviruses and have been implicated in the emergence of several genotypes, including endemic human coronaviruses [3-6]. The involvement of genetic recombination in the origin of SARS-CoV-2 is also suspected [7]. Regarding SARS-CoV-2, coinfection in the same patient with distinct variants has been reported [8-14]. In addition, several studies have described or suspected genetic recombinations for this virus [10,13-14,15-25]. However, most of these recombinants relied solely on the coexistence of signature mutations of different SARS-CoV-2 variants in ge-nomes obtained from a single patient’s sample, and they were not isolated in culture. Since January 2020, our laboratory has screened more than one million respiratory specimens for SARS-CoV-2 infection by real-time reverse transcription-PCR (qPCR) without interruption or limited capacity including for all patients sampled in our institute and in the Marseille public hospitals [26,27]. This has provided us with sequential samples from multiple patients, and enabled us to detect reinfections, and prolonged or even chronic infections in severely immunocompromised patients [28-30]. Here, we re-port the gradual creation of a recombinant SARS-CoV-2 involving two variants in a lymphoma patient chronically infected over a period of 14 months, and the isolation of the recombinant virus in culture.

## RESULTS

### Chronic SARS-CoV-2-infection in a severely immunocompromised patient

An immunocompromised adult patient had an uncontrolled SARS-CoV-2 infection for 14 months until death (Supplementary Material: Supplementary Methods and Results). This patient had been previously diagnosed with mixed Hodgkin and follicular lymphoma. In 2020, the patient developed severe SARS-CoV-2-associated pneumonia as diagnosed virologically by qPCR. Despite clinical improvement, viral clearance did not occur as SARS-CoV-2 RNA remained detectable by qPCR on most nasopharyngeal samples collected until patient’s death. qPCR was negative at Month 5 of diagnosis but positive when re-tested at Month 7, and then only transiently negative for ≤3 days.

### Evidence of hybrids of variants

After 14 months of infection, we identified a virus whose genome was a hybrid of two known variants, B.1.160 [according to Pangolin lineage (https://cov-lineages.org/resources/pangolin.html) [31]] [a.k.a. Nextstrain clade (https://nextstrain.org/) [32] 20A.EU2, or Marseille-4 [27]] and Alpha [according to the WHO denomination (https://www.who.int/fr/activities/tracking-SARS-CoV-2-variants) (a.k.a. 20I or B.1.1.7)] (Figure 1; Figure 2).

**Figure 1.**
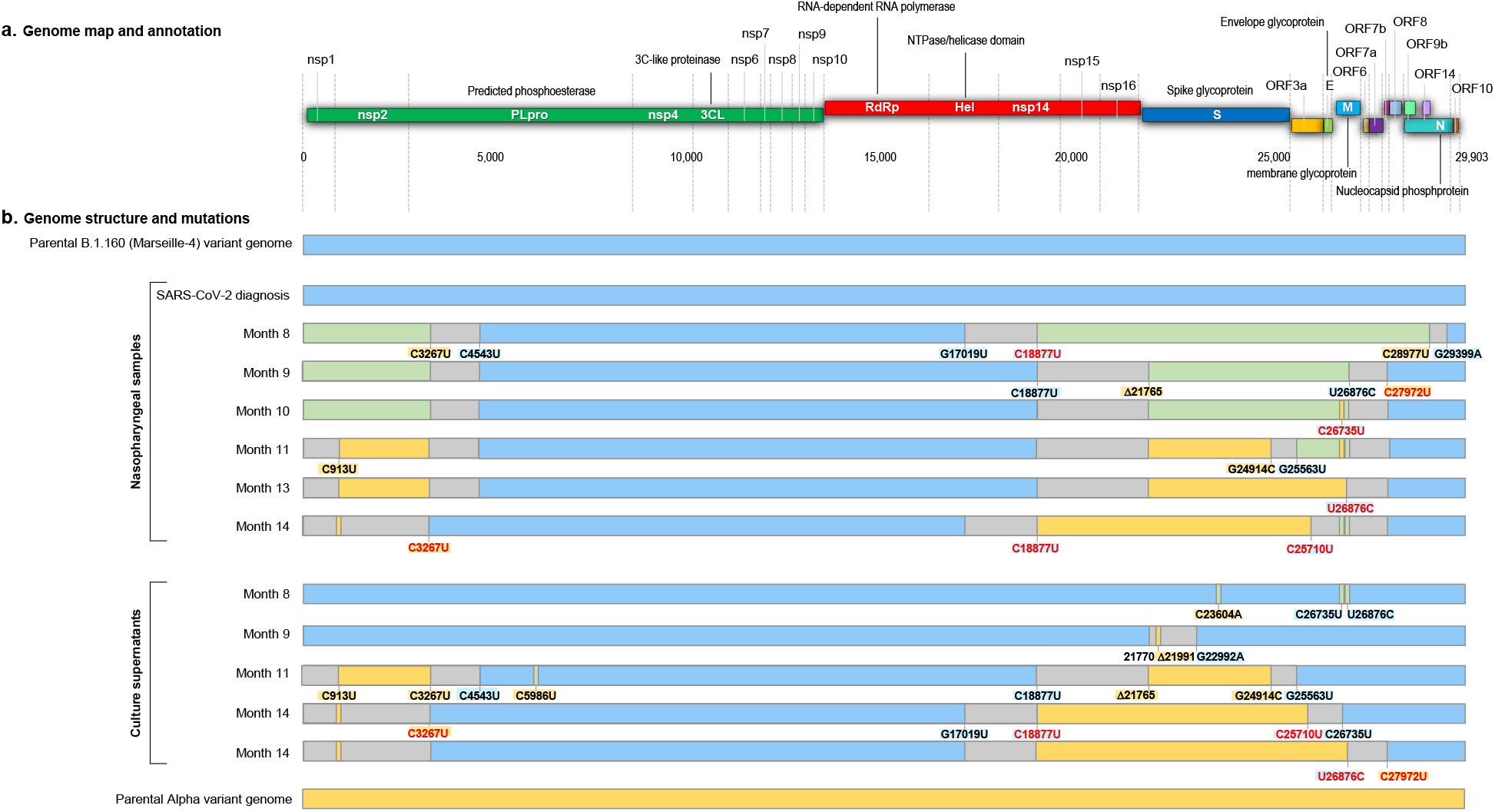
Schematic representation of the structure of the SARS-CoV-2 genomes obtained from the nasopharyngeal samples and from the culture supernatants and of the recombination events over time, in reference to parental genomes of the B.1.160 and Alpha variants. a. Genome map and annotation. b. Genome structure and mutations. Blue color of rectangles indicates sequences from a B.1.160 variant; yellow color indicates sequences from an Alpha variant; green color indicates co-detection of sequences from a B.1.160 variant and from an Alpha variant; grey color indicates sequences from indeterminate origin. Signature mutations from the B.1.160 and Alpha variants are indicated by a blue background and a yellow background, respectively. Signature mutations that are absent are indicated by a red font. Δ21765: -6 nucleotides; Δ21991: -3 nucleotides. Nsp, nonstructural protein; ORF, open reading frame.

**Figure 2.**
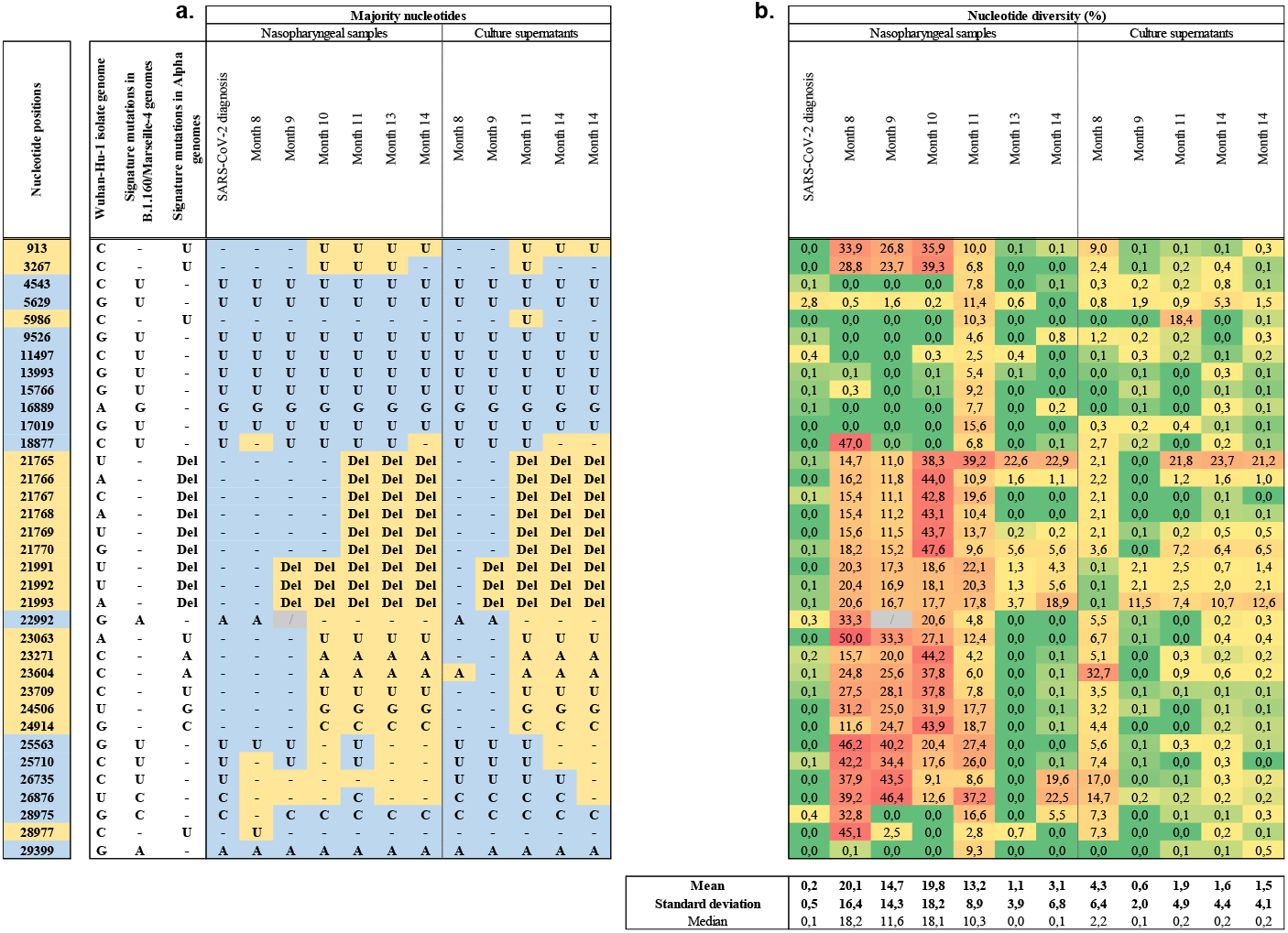
Majority nucleotides (a) and nucleotide diversity (b) for sequences obtained from the respiratory samples and the culture supernatant at nucleotide positions of the SARS-CoV-2 genome that harbor signature mutations of the B.1.160 or Alpha variants. Nucleotide positions are in reference to the genome of the Wuhan-Hu-1 isolate GenBank accession no. NC_045512.2. a: B.1.160 nucleotides are indicated by a blue background; Alpha nucleotides are indicated by a yellow background. Del, nucleotide deletion b: Nucleotide diversity is the proportion of sequence reads that do not harbor the consensus (majority) nucleotide.

This hybrid genome was obtained from the respiratory samples by next-generation sequencing, as previously described [27]. In addition, the hybrid virus was isolated in culture, as previously described [33]. The hybrid genome sequence consisted of a B.1.160 variant matrix, of which two regions, the first one being located at the 5’ tip of the ge-nome and containing synonymous mutation C913U and the second one spanning from positions 17,109-18,877 to positions 25,710-27,972, were replaced by those of an Alpha variant (Figure 1; Figure 2; Supplementary Material: Figure S1). All eight signature mu-tations of the Alpha variant were detected in the spike gene in the absence of the S477N mutation that is a signature of the B.1.160 variant. Nucleotide diversity at the 35 positions harboring signature mutations of the Alpha or B.1.160 variants was low [mean (±standard deviation) value of 3.1±6.8%] (Figure 2; Supplementary Material: Figure S1), indicating that the hybrid content of the genome was not explained by a co-infection by the two variants or by contamination. These findings indicated that this mosaic genome was the result of recombinations between parental genomes of the B.1.160 and Alpha variants.

### Genome of the initial virus

By analyzing the sequential samples available from this patient, we were able to determine that he was first infected with the B.1.160 variant, which was epidemic at the time of diagnosis of his infection during summer 2020. This variant predominated in our region from August 2020 until January 2021 and was replaced by the Alpha variant that emerged in December 2020 [27]. SARS-CoV-2 could not be isolated retrospectively from this sample, but its genome was typical of a B.1.160 variant and displayed no significant nucleotide diversity (mean, 0.2±0.5%). It was classified by phylogeny as a B.1.160 variant (Figure 3). Unfortunately, although SARS-CoV-2 qPCR was still positive in another laboratory at Month 4 post-diagnosis, the sample was unavailable, and we were not able to verify that only this virus had persisted and that there was no co-infection with an Alpha variant at this time. Thus, we did not obtain the genome and an isolate of the Alpha variant. The closest sample in time to the initial one dated from Month 8, and it already demonstrated a mosaicism between genomes of B.1.160 and Alpha variants (Figure 1, Figure 2).

**Figure 3.**
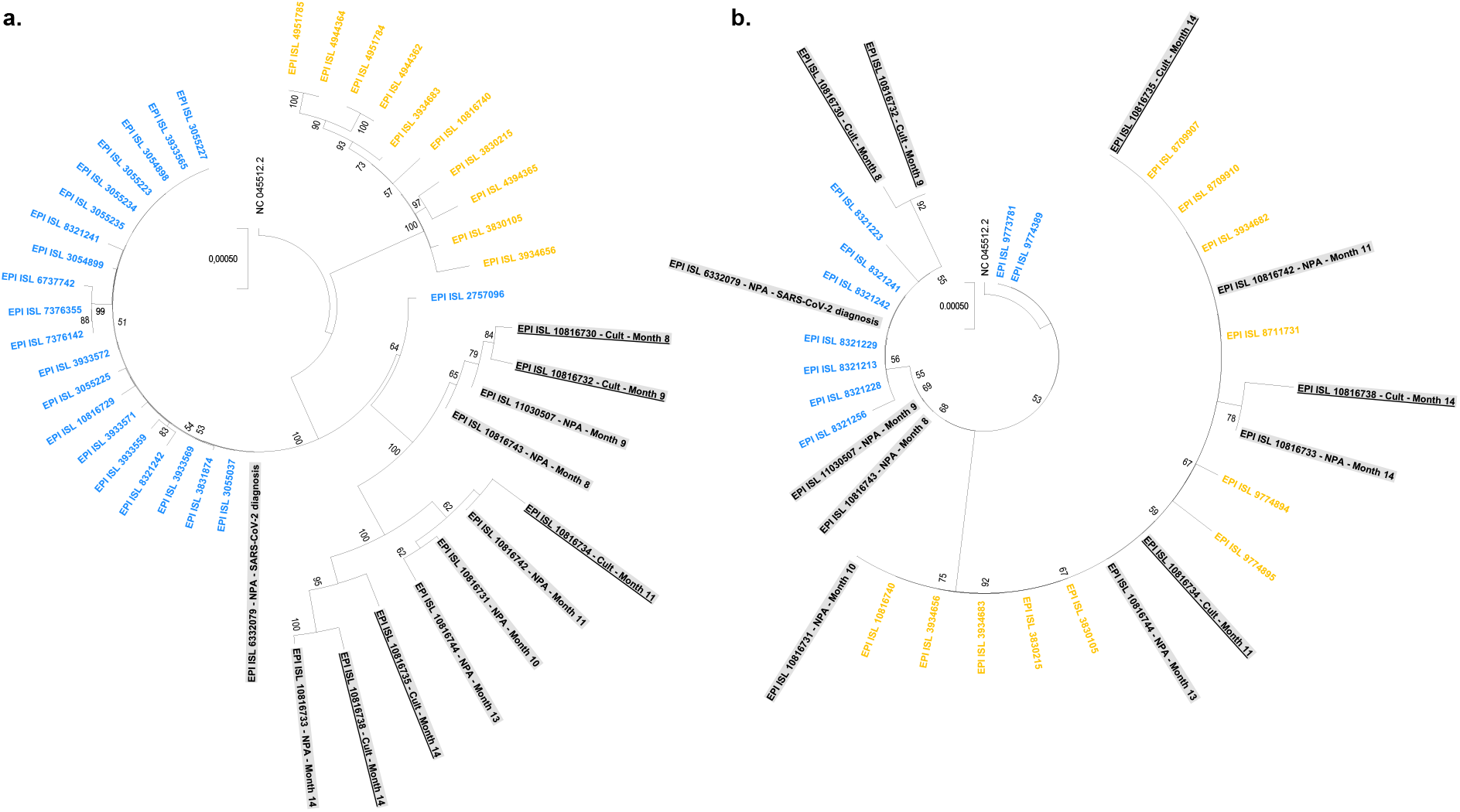
Phylogenetic analyses based on SARS-CoV-2 genomes (a) and spike gene sequences (b). Sequences obtained from the case-patient are indicated by a grey background, and those obtained from cultures are underlined. Other sequences from our SARS-CoV-2 sequence database are indicated by a blue font when classified as of the B.1.160 variant, and by a yellow font when classified as of the Alpha variant. Sequences are labeled with their GISAID (https://www.gisaid.org/) [34] identifiers. Trees are rooted with the genome of the Wuhan-Hu-1 isolate GenBank accession no. NC_045512.2.

### Steps in creation of the recombinant

We used three procedures to characterize the different recombination steps by analyzing seven sequential respiratory samples collected from the patient (Tables 1, 2). First, sequencing from the respiratory samples of the viral genomes; second, sequencing from the respiratory samples of PCR products overlapping the putative recombination sites; and third, viral culture with sequencing of the genomes of the isolates. These approaches allowed us to evidence that several viruses and recombinant forms had coexisted in the sequential samples, as signature mutations of the two variants were co-detected at multiple positions, with a nucleotide diversity that reached high levels and evolved over time (Figure 2; Supplementary Material: Supplementary Results and Figure S1). We observed an evolution towards the genome sequence of the recombinant virus that predominated at the end of the patient’s follow-up, following recombination events at three sites between parental genomes of B.1.160 and Alpha variants, with a low level of nucleotide diversity observed at that time at the positions harboring signature mutations of these variants.

**Table 1.** Genome sequences obtained from the sequential nasopharyngeal samples of the case-patient

| GISAID identifier | Sampling time post-diagnosis (months) | Next-generation sequencing technology, instrument |
| --- | --- | --- |
| EPI_ISL_6332079 | M0 | Illumina, NovaSeq |
| EPI_ISL_10816743 | Month 8 | Illumina, NovaSeq |
| EPI_ISL_11030507 | Month 9 | Illumina, NovaSeq |
| EPI_ISL_10816731 | Month 10 | Illumina, NovaSeq |
| EPI_ISL_10816742 | Month 11 | Nanopore, GridION |
| EPI_ISL_10816744 | Month 13 | Illumina, NovaSeq |
| EPI_ISL_10816733 | Month 14 | Illumina, NovaSeq |

**Table 2.**
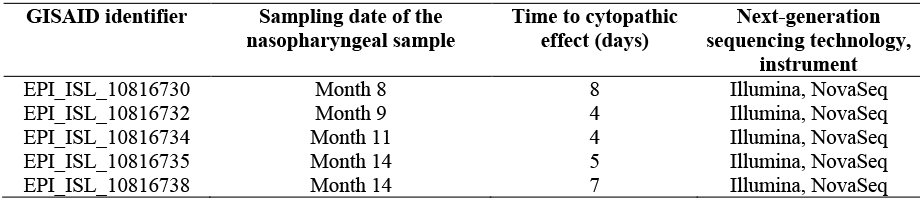
Genome sequences obtained from the culture supernatants

| GISAID identifier | Sampling date of the nasopharyngeal sample | Time to cytopathic effect (days) | Next-generation sequencing technology, instrument |
| --- | --- | --- | --- |
| EPI_ISL_10816730 | Month 8 | 8 | Illumina, NovaSeq |
| EPI_ISL_10816732 | Month 9 | 4 | Illumina, NovaSeq |
| EPI_ISL_10816734 | Month 11 | 4 | Illumina, NovaSeq |
| EPI_ISL_10816735 | Month 14 | 5 | Illumina, NovaSeq |
| EPI_ISL_10816738 | Month 14 | 7 | Illumina, NovaSeq |

## DISCUSSION

We highlight here in an immunocompromised lymphoma patient who received several treatments the presence after 14 months of infection of a hybrid virus of two known variants, B.1.160 and Alpha, which successively predominated in our region during the period during which this patient was followed-up [35, 27]. The first evidence of a hybrid genome was obtained 8 months after the diagnosis of infection by the B.1.160 variant, and the absence of available samples covering this period did not allow us to date the superinfection by the Alpha variant. The signature mutations of the Alpha variant observed in the hybrid genomes between 8 and 14 months cannot have occurred randomly considering their number and distribution along the genome and their majority presence, and their location indicates recombinations in three regions. In addition, genomic analyses carried out for sequential respiratory samples and viral cultures demonstrate the successive presence of several viruses with hybrid genomes, one of them having established itself in this patient and remaining the one that continued circulating until his death.

We believe that this observation, which sheds light on the recombination mecha-nism of RNA viruses, is significant, and to our knowledge this is the first one describing, through the analysis of sequential samples over more than a year, the creation of a re-combinant SARS-CoV-2 and the isolation of the recombinant virus in culture. Sixteen interlineage recombinants between the Alpha variant and non-Alpha viruses were reported in 2021 in the UK, of 279,000 genomes analyzed [8]. In addition, 1,175 (0.2%) pu-tative recombinant genomes were identified among 537,360 genomes, and it was reported that up to 5% of SARS-CoV-2 that circulated in the USA and UK might be recombinants [18]. Moreover, the number of cases of detection of recombinant genomes is growing [10,13-14,15-25]. Recently, coronaviruses have been reported to be able to integrate se-quences putatively from insects [36]. Such natural mosaicisms in these viruses make it possible to understand the emergence of RNA viruses and should lead to a strengthening of genomic surveillance in patients presenting with coinfections by several RNA viruses, as is observed in patients infected with several respiratory viruses including SARS-CoV-2, endemic human coronaviruses, influenza viruses, or rhinoviruses [37]. Such infectious episodes could perhaps lead to the creation of new emerging viruses, as has been for instance reported for enteroviruses of humans and great apes [38].

## MATERIALS AND METHODS

### SARS-CoV-2 genome sequencing

SARS-CoV-2 genome sequencing was performed as previously described. Briefly, viral RNA was extracted from 200 μL of nasopharyngeal swab fluid using the EZ1 Virus Mini kit v2.0 on an EZ1 Advanced XL instrument (Qiagen, Courtaboeuf, France) or using the MagMax Viral/Pathogen Nucleic Acid Isolation kit on the KingFisher Flex system (Thermo Fisher Scientific, Waltham, MA, United States), following the manufacturer’s instructions.

SARS-CoV-2 genome sequences were obtained by next-generation sequencing with various procedures with the Illumina COVIDSeq protocol on a NovaSeq 6000 instrument (Illumina Inc.), or by multiplex PCR with ARTIC nCoV-2019 V3 Panel primers (IDT, Coralville, IA, USA) combined with the Oxford Nanopore technology (ONT) on a GridION instrument (Oxford Nanopore Technologies Ltd., Oxford, United Kingdom), as previously described [27,14]. After its extraction, viral RNA was reverse-transcribed according to the COVIDSeq protocol (Illumina Inc.) or using the LunaScript RT SuperMix kit (New England Biolabs) when performing next-generation sequencing with the Nanopore technology, following the manufacturer’s recommendations.

### Genome analysis

After using the Nanopore technology combined with the ARTIC protocol, fastq files were processed using the ARTIC field bioinformatics pipeline (ARTIC-nCoV-bioinformaticsSOP-v1.1.0; https://github.com/artic-network/fieldbioinformatics), as previously described [27,14]. NGS reads were basecalled using Guppy (4.0.14) and aligned to the genome of the Wuhan-Hu-1isolate, GenBank accession No. MN908947.3, using minimap2 (v2.17-r941) (https://github.com/lh3/minimap2) [39]. The ARTIC tool align_trim was used to softmask primers from read alignment and cap sequencing depth at a maximum of 400-fold coverage. Consensus-level variant candidates were identified using a threshold of 70% and the Medaka (v.0.11.5) workflow developed by ARTIC (https://github.com/artic-network/artic-ncov2019). From the unique sequence obtained with the ARTIC-Nanopore technology, a sorted bam file was loaded on the CLC Genomics workbench v7 software and a tsv file was then exported. NovaSeq reads were basecalled using the Dragen Bcl Convert pipeline (v3.9.3; https://emea.support.illumina.com/sequencing/sequencing_software/bcl-convert.html (Illumina Inc.)). Mapping was carried out on the Wuhan-Hu-1 isolate genome with the bwa-mem2 tool (v2.2.1; https://github.com/bwa-mem2/bwa-mem2) and was cleaned with Samtools (v1.13; https://www.htslib.org/) [40]. Variant calling was performed with freebayes (v1.3.5; https://github.com/freebayes/freebayes) [41] and consensus genomes were built with Bcftools (v1.13; https://samtools.github.io/bcftools/bcftools.html). Freebayes results were filtered with a threshold of 70% for the majority nucleotide. A tsv file was generated using an in-house Python script. The clade was designated at the consensus level with the Nextclade online tool (https://clades.nextstrain.org/) [42,32] and an in-house Python script allowed detection of variants and hybrids of variants. At the sub-consensus level, variant frequencies compared to the Wuhan-Hu-1 isolate genome were calculated from tsv files. Nucleotide diversity at genomic positions was calculated using the Microsoft Excel software (https://www.microsoft.com/en-us/microsoft-365/excel) with an in-house built file. It corresponded to the proportion of sequence reads that do not harbor the consensus (majority) nucleotide. Genome sequences obtained in the present study were submitted to the GISAID sequence database (https://www.gisaid.org/) [34] (see Supplementary Material: Supplementary Table S1).

### Generation of additional sequence reads

Sequencing of reverse-transcription-PCR-targeted regions: Extraction of the RNA samples was carried out using the EZ1 Virus kit with the EZ1 Advanced XL instrument (Qiagen) following the manufacturer’s recommendations. PCR amplification of the 3 regions was amplified in a 25 μL total volume using the SuperScript III One-Step RT-PCR Kit (Invitrogen, Carlsbad, CA, USA), using primer concentrations of 200 nM per reaction. PCR were performed with following conditions: 50°C for 25 min, 95°C for 2 min, then 40 cycles including 15 s at 95°C, 45 s at 60°C, and 2 min at 68°C. Sequences of PCR primers are provided in the Supplementary Material. Amplicons were sequenced with Nanopore technology on a GridION instrument (Oxford Nanopore Technologies Ltd.), following manufacturer’s instructions. Fastq files were processed as described above. Continuous reads overlapping signature mutations of distinct variants were filtered using SAMtools (v1.13; https://github.com/samtools/) [40] combined with an in-house awk script. Reads were then filtered according to variant-specific nucleotide patterns using SAMtools combined with an in-house awk script. Groups of reads with same patterns of mutations were then visualized using the IGV software (https://software.broadinstitute.org/software/igv/) [43].

Metagenomic sequencing: Nucleic acid extraction was performed with the EZ1 Virus kit with the EZ1 Advanced XL instrument (Qiagen) following the manufacturer’s recommendations, using 200 µL of sample and eluting in 60 µL of elution buffer. Reverse transcription was performed with all 60 µL of this solution using the TaqMan Reverse Transcription Reagent kit (Applied-Biosystems, Foster City, CA, USA), according to the manufacturer’s protocol under the following conditions: 10 min at 25°C, 30 min at 48°C, and 5 min at 95°C. Then, 300 µL of obtained cDNA was transferred in a 1.5 mL Eppendorf LoBind tube (Eppendorf, Le Pecq, France). Second DNA strand was synthesized by adding a mix of 24 µL of Klenow Fragment DNA polymerase (New England Biolabs, Beverly, MA, USA), 66 µL of nuclease-free water, 45 µL of NEB Buffer 2 (New England Biolabs), and 15 µL of dNTPs working solution produced with 10 µL of each dNTP at a 100 mM concentration, and 60 µL of nuclease-free water (New England Biolabs). This mix was kept at 37°C for one hour. A purification step consisted in adding 450 µL of magnetic CleanNGS beads for a 1:1 volume ratio (CleanNA, Waddinxveen, the Netherlands) then incubating for 5 min in a magnetic support, and washing with 1,000 µL of ethanol at 70%, before elution of beads in 50 µL of Tris EDTA 1X with centrifugation for 10 min at 300 rpm at room temperature. Subsequently, DNA library was prepared with the DNA ligation sequencing kit SQK-LSK109 (Oxford Nanopore Technologies Ltd.), and next-generation sequencing was performed with the Nanopore technology on a PromethION instrument (Oxford Nanopore Technologies Ltd.). Each sample was sequenced on a different PromethION Flow cell R10.4 (Oxford Nanopore Technologies Ltd.).

### Phylogenetic analysis on whole and partial genomes

Phylogenetic analyses were performed separately for the twelve genome sequences and the twelve spike gene sequences obtained from the nasopharyngeal samples or the culture supernatants. Sequences were aligned using MAFFT v.7 [44] with their 20 most similar hits identified with the BLAST tool [45] among SARS-CoV-2 genomes from our database that contains sequences obtained from clinical samples collected between February 2020 and February 2022 [27,14]. Phylogeny reconstruction was performed using the IQ-TREE software with the GTR Model and 1,000 ultrafast bootstrap repetitions (http://www.iqtree.org) [46], and trees were visualized with iTOL (Interactive Tree Of Life) (https://itol.embl.de/) [47] and MEGA X (v10.2.6; https://www.megasoftware.net/) [47] softwares.

### Virus culture isolation

Culture isolation was performed on Vero E6 cells, as previously described [33].

## Data Availability

The dataset generated and analyzed during the current study is available in the GISAID sequence database (https://www.gisaid.org/).

## Acknowledgments

We are thankful to Ludivine Brechard, Claudia Andrieu, Raphael Tola, Jeremy Delerce for their technical help; to Sophie Amrane and Justine Raclot for their help with the clinical management; and to the medical biology laboratory Alphabio, hôpital Européen, Marseille, France, for providing some results of SARS-CoV-2 qPCR testing.

## Author contributions

P.C., B.L.S., and D.R. conceived the project. E.B., P.C., J.-C.L., A.L., M.B., and P.L. provided materials or performed analyses. E.B., P.C., J.-C.L., A.L., M.B., P.-E.F., B.L.S., and D.R. analyzed the data. E.B., P.C., P.-E.F., B.L.S, and D.R. drafted the paper. All authors participated in the discussion and interpretation of the results. All authors edited and proofread the final manuscript. All authors have read and agreed to the published version of the manuscript.

## Data availability

The dataset generated and analyzed during the current study is available in the GISAID sequence database (https://www.gisaid.org/) [34].

## Ethics

This study has been approved by the ethics committee of University Hospital Institute (IHU) Méditerranée Infection (No. 2022-008). Access to the patients’ biological and registry data issued from the hospital information system was approved by the data protection committee of Assistance Publique-Hôpitaux de Marseille (APHM) and was recorded in the European General Data Protection Regulation registry under number RGPD/APHM 2019-73.

## SUPPLEMENTARY MATERIAL

### SUPPLEMENTARY METHODS

At our institute, SARS-CoV-2 real-time reverse-transcription-PCR (qPCR) was performed on nasopharyngeal samples collected from patients tested by real-time reverse-transcription-PCR (qPCR) using the BGI real-time fluorescent RT-PCR assay (BGI Genomics, Shanghai Fosun Long March Medical Science Co., Ltd., Shenzhen, China) or the NeuMoDx SARS-CoV-2 assay (NeuMoDx Molecular, Ann Arbor, Michigan). Outside our institute (medical biology laboratory Alphabio, hôpital Européen, France), SARS-CoV-2 qPCR was performed using the BD MAX System (Becton Dickinson, Sparks, MD, USA).

### SUPPLEMENTARY RESULTS

#### Virological follow-up by SARS-CoV-2 real-time reverse-transcription-PCR (qPCR)

SARS-CoV-2 qPCR performed on nasopharyngeal samples was positive between summer 2020 (at time of SARS-CoV-2 diagnosis) and Month 4 post-diagnosis, then negative once at Month 5 but positive again on the next sample tested in Month 7. Between Month 8 and Month 11, qPCR remained almost always positive being only transiently negative for ≤3 days. Then qPCR was consistently positive when performed between Month 13 and Month 14 post-diagnosis.

#### Nucleotide diversity at positions harboring signature mutations of the B.1.160 or Alpha variants

Regarding the SARS-CoV-2 genomes obtained from the respiratory samples, which were obtained with the Illumina technology except for the sample collected at Month 11 post-SARS-CoV-2 diagnosis for which the genome was obtained with the Nanopore technology, nucleotide di-versity at the 35 positions harboring signature mutations of the B.1.160 or Alpha variants differed according to the time of sample collection. It was low at time of diagnosis in 2020, with a mean (±standard deviation) value of 0.2±0.5%, increased to reach high values at Month 8 (20.1±16.4%), at Month 9 (14.7±14.3%), at Month 10 (19.8±18.2%), and at Month 11 (13.2±8.9%), then decreased to low values again at Month 13 (1.1±3.9%) and at Month 14 (3.1±6.8%) (Figure 2 of the main text). The low nucleotide diversity observed for genomes obtained at Month 13 and at Month 14 indicated that the mosaicism consisting in the presence of signature mutations of the two variants was not explained by a co-infection by these two variants or by a contamination of the samples prior or during the next-generation sequencing procedure.

Regarding the genomes obtained from the culture supernatants, which were all ob-tained with the Illumina technology, nucleotide diversity at positions harboring signature mutations of the B.1.160 or Alpha variants was low, indicating that these genomes, including those recombinants, were generated from a single viral isolate. Indeed, mean nucleotide diversity ranged between 0.6±2.0% and 4.3±6.4% (Figure 2 of the main text). The viral genomes obtained from the culture of the respiratory samples collected at Month 14 were highly similar to the viral genome obtained directly from the respiratory sample collected at Month 14.

#### Generation of additional sequence reads

Sequencing of reverse-transcription-PCR-targeted regions: We attempted to generate sequence reads that are hybrids of the B.1.160 and Alpha variants from the respiratory sample collected at Month 11 by PCR amplification of the regions overlapping re-combination sites then next-generation sequencing with the Oxford Nanopore Technol-ogy (ONT) on a GridION instrument (Oxford Nanopore Technologies Ltd., Oxford, United Kingdom).

Amplicons corresponding to positions 3,100-4,570 (in reference to the genome of the Wuhan-Hu-1 isolate GenBank accession no. NC_045512.2) were amplified using the following primers (in 5’-3’ orientation): Forward: TTCAACCTGAAGAA-GAGCAA; reverse: TGGCATTGTAACAAGAGTTT. Amplicons corresponding to posi-tions 24,813-29,074 were amplified using the following primers: Forward: ATGGAAAAGCACACTTTCCT; reverse: GCTTTAGTGGCAGTACGTTT. For the am-plicons corresponding to positions 3,100-4,570, 87% of the reads were hybrids harboring both Alpha (C3267U) and B.1.160 (C4543U) signature mutations. For the amplicon cor-responding to positions 24,813-29,074, 70% of the reads were hybrids harboring Alpha (G24914C) and B.1.160 (G25563U, C25710U, C26735U, U26876C and G28975C) mutations, and 24% were hybrids harboring Alpha mutation G24914C and B.1.160 mutation G28975C.

Metagenomic sequencing: Next-generation sequencing was also carried out using a metagenomic approach with the Nanopore technology on a GridION instrument (Oxford Nanopore Technologies Ltd.) to attempt obtaining long sequence reads and detect addi-tional reads that are hybrids of the B.1.160 and Alpha variants. For the first recombination site, four reads with a length ranging between 1,855-11,979 nucleotides were obtained, which harbored signature mutations of the Alpha (C3267U) and B.1.160 (C4543U) variants. For the third recombination site, four reads with a length comprised between 1,486-8,143 nucleotides were obtained that harbored signature mutations of the Alpha (G24914C) and B.1.160 (C25563U) variants.

#### Viral culture

Cytopathic effects were observed between 4 and 8 days after the inoculation of the nasopharyngeal samples on Vero E6 cells as previously described [1] (Table 2 of the main text).

#### Phylogenetic analyses based on SARS-CoV-2 genomes and spike gene sequences

Phylogeny reconstructions were performed based on viral genomes or spike genes, which included sequences obtained directly from the nasopharyngeal samples and from the cultures, and their best BLAST hits from the IHU Méditerranée Infection sequence database that were classified as B.1.160 or Alpha variants [2]. The genome obtained from the nasopharyngeal sample collected in 2020 at time of SARS-CoV-2 diagnosis was clustered with genomes of the B.1.160 variant, apart from the other genomes obtained from the case-patient (Figure 3a of the main text). Other genomes obtained from the nasopharyngeal samples collected since Month 8 and those obtained from cultures of the nasopharyngeal samples collected since Month 8 were also clustered with genomes of the B.1.160 variant, but they were clustered together, apart from the other genomes. The Alpha variant genomes were clustered apart from all other genomes. Spike gene phylogeny showed the clusterisation with sequences of the B.1.160 variant of the sequences obtained from the case-patient either directly or post-culture from samples collected between SARS-CoV-2 diagnosis and Month 9 post-diagnosis (Figure 3b of the main text). In contrast, sequences obtained from the case-patient either directly or post-culture from samples collected since Month 10 post-diagnosis were clustered with Alpha variant sequences.

### SUPPLEMENTARY FIGURES

**Supplementary Figure S1.**
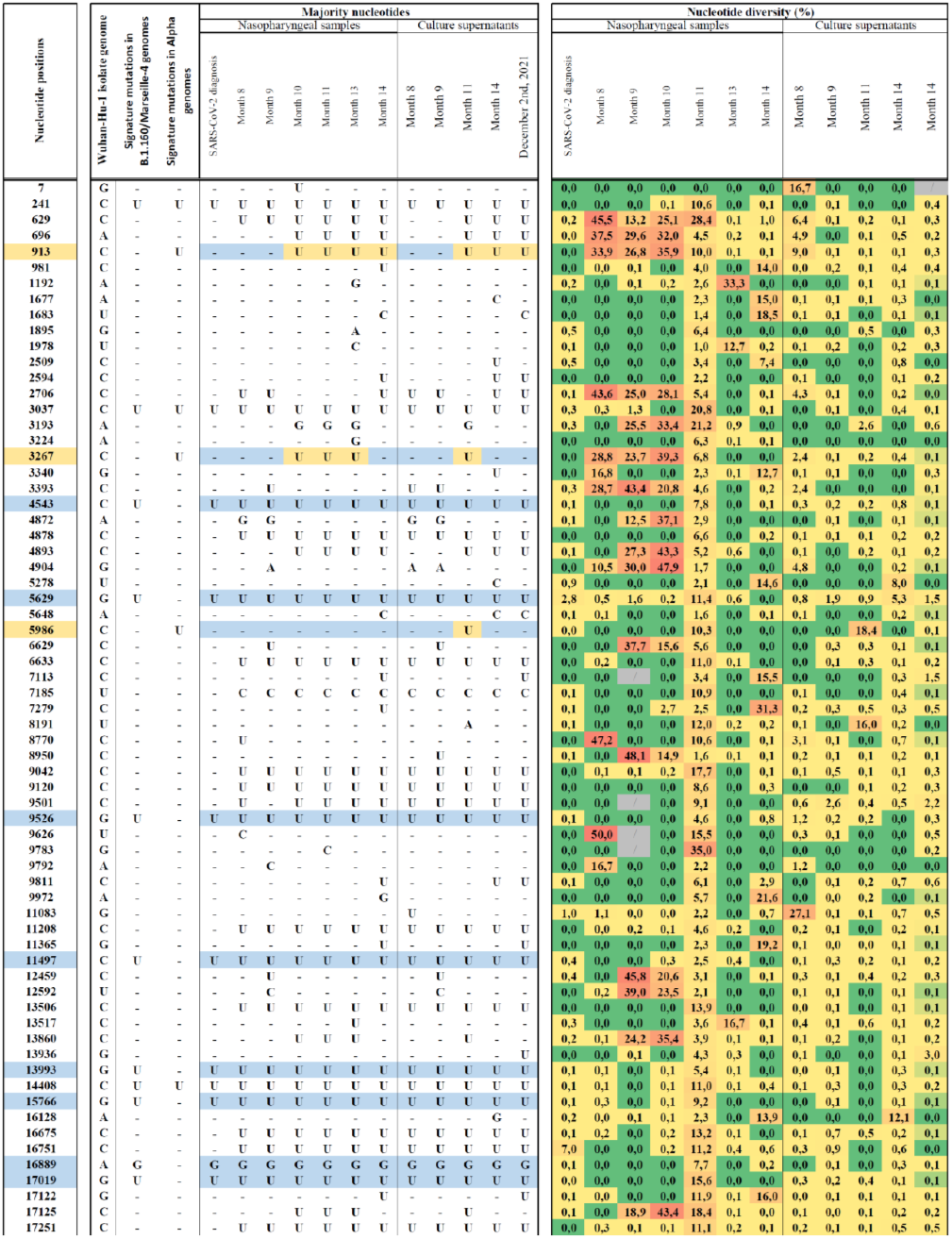
**Majority nucleotides and nucleotide diversity for sequences obtained from the respiratory samples and the culture supernatant at nucleotide positions of the SARS-CoV-2 genome that harbor signature mutations of the B.1.160 or Alpha variants and at any other positions that harbor mutations.** Nucleotide positions are in reference to the genome of the Wuhan-Hu-1 isolate GenBank accession no. NC_045512.2. B.1.160 nucleotides are indicated by a blue background; Alpha nucleotides are indicated by a yellow background. Del, nucleotide deletion. Nucleotide diversity is the proportion of sequence reads that do not harbor the consensus (majority) nucleotide.

**Supplementary Figure S2.**
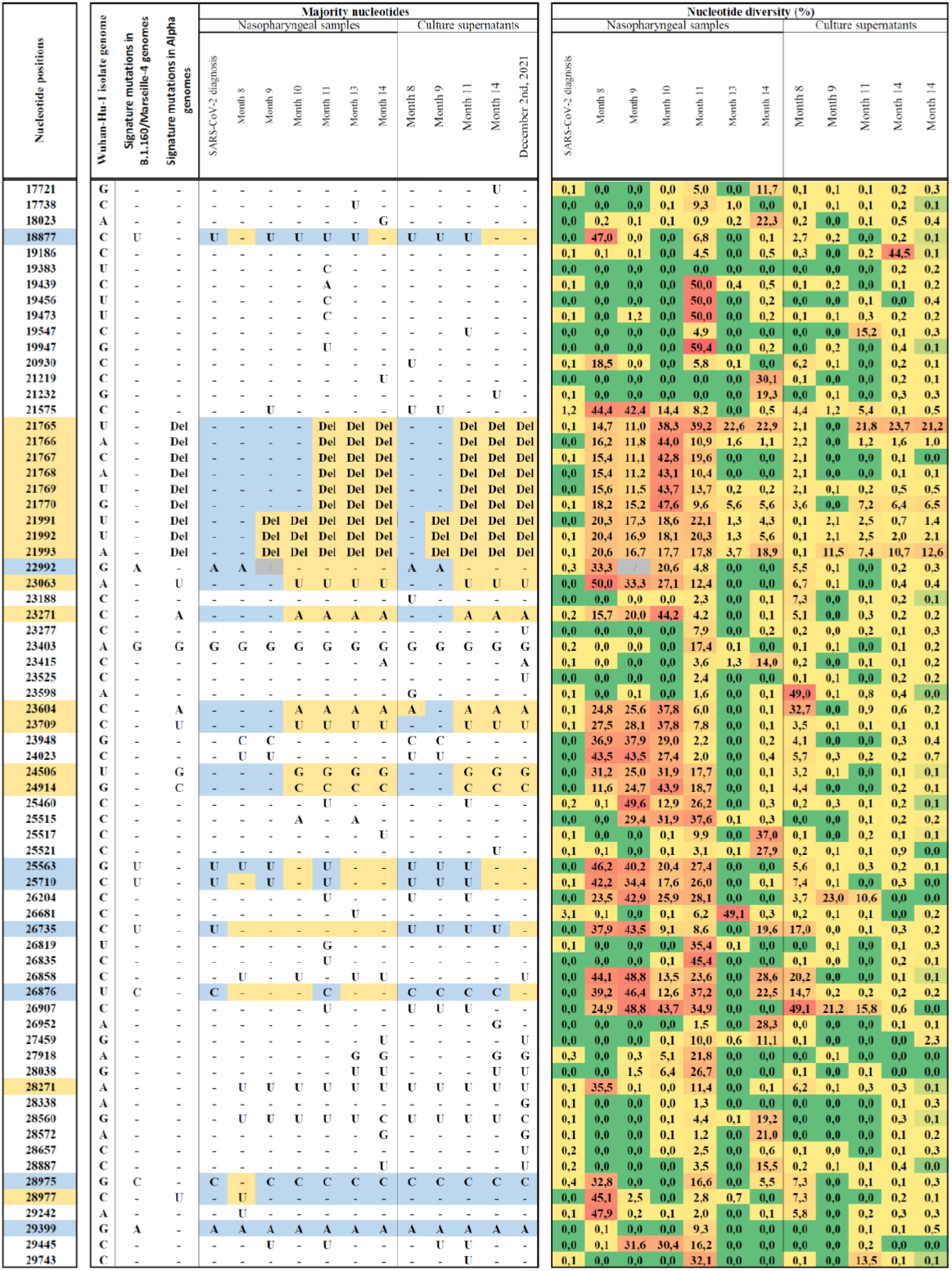

### SUPPLEMENTARY TABLES

**Supplementary Table S1.**
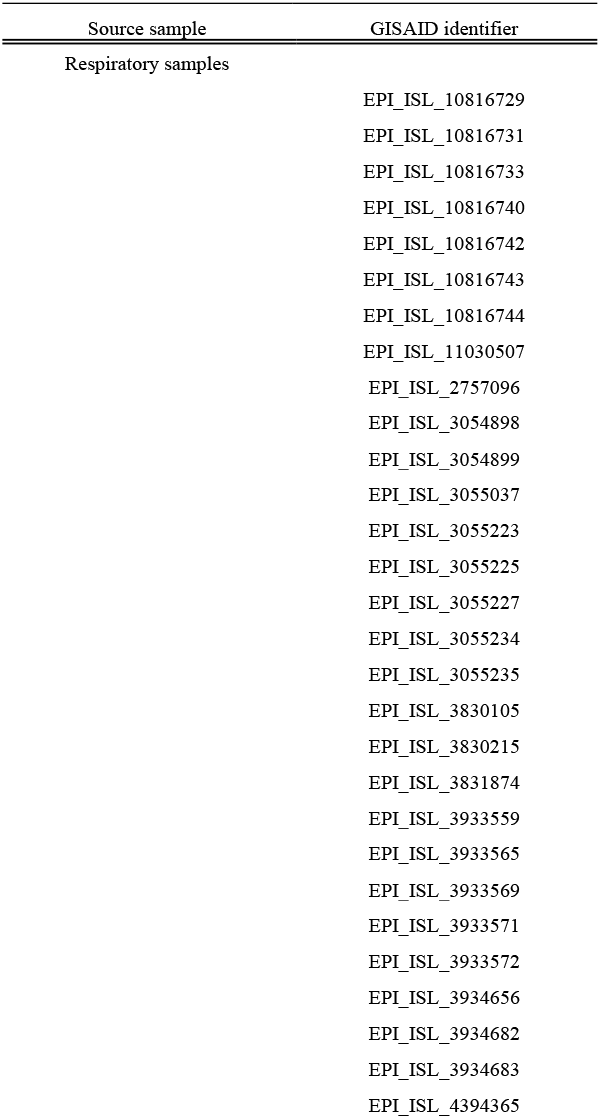
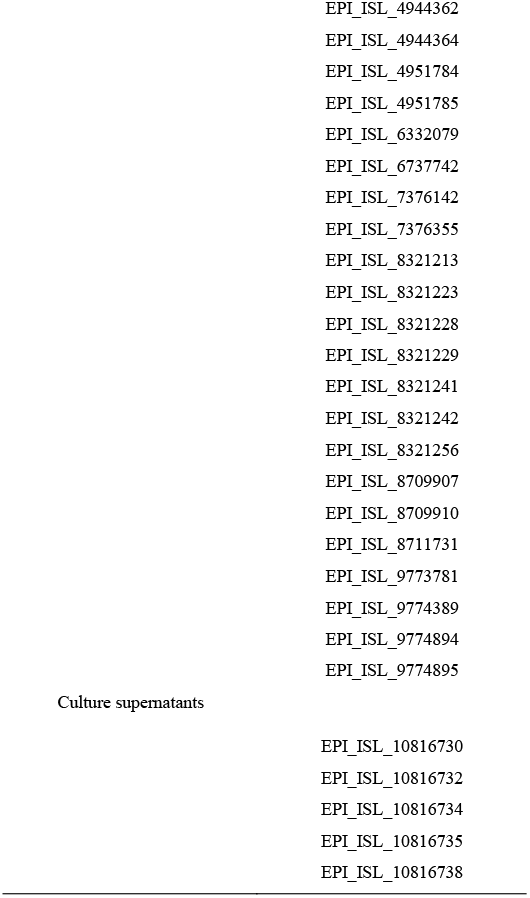
List of GISAID identifiers for sequences used in the present study. All genomes analyzed here were obtained in our laboratory (University hospital institute Méditerranée Infection, Marseille, France). The GISAID sequence database is accessible at: (https://www.gisaid.org/) [3].

